# The Association Between Oral Microbiota and Chronic Obstructive Pulmonary Disease: An Integrated Study of Genetic Causal Inference and Bioinformatics Analysis

**DOI:** 10.64898/2025.12.01.25341371

**Authors:** Xiao-Jie An, Zi-Feng Wei, Yi-Ting Huang, Jia-Pei Wuzhang, Xin-Xin Zhang, Hao-Yu Li, Run-Ze Liu

## Abstract

**Background:** Chronic obstructive pulmonary disease (COPD) is the third leading cause of global mortality. Emerging evidence suggests the oral microbiome may contribute to COPD progression, though causal relationships remain elusive.

**Methods:** Using bidirectional Mendelian randomization (MR) on East Asian genome-wide association study (GWAS) summary data, we assessed causal links between oral microbial taxa and COPD risk. Subsequently, hub genes in COPD bulk RNA sequencing data were identified by integrating the Protein-Protein Interaction (PPI) network with logistic regression, followed by target validation using single-cell RNA sequencing, immune infiltration analysis, and molecular docking.

**Results:** Forward MR identified 48 taxa associated with COPD, primarily from genera such as *Fusobacterium*, *Prevotella*, and *Streptococcus*. Reverse MR detected 79 taxa affected by COPD, mainly involving *Campylobacter*, *Rothia*, and *Streptococcus*. Through the PPI network, logistic regression screening, and multi-omics analysis validation, *MPDZ* emerged as a key hub gene, upregulated in Ciliated cells and linked to immune dysregulation. Molecular docking revealed six candidate drugs with strong binding affinity to *MPDZ*.

**Conclusion:** Our study identified bidirectional causal associations between the oral microbiota and COPD in East Asian populations and prioritized candidate targets relevant to COPD pathogenesis, potentially offering new insights for future mechanistic studies and therapeutic exploration.

## 1. Introduction

COPD ranks as the third leading cause of death worldwide, characterized primarily by airflow limitation and structural lung damage driven by chronic inflammation of the airways and pulmonary parenchyma [1]. The disease is driven by chronic inflammatory responses that cause airway remodeling and alveolar wall destruction, clinically manifesting as chronic cough, expectoration, and progressive dyspnea [2]. Recent global epidemiological data indicate that the overall burden of COPD has been increasing since 1990, with rising prevalence and mortality rates observed annually [3, 4]. Environmental exposures, genetic susceptibility, pulmonary inflammation, and oxidative stress are well-recognized mechanisms contributing to COPD pathogenesis [5]. However, these pathways only partially explain the complex etiology of COPD. Current research has yet to fully elucidate its pathophysiology or yield truly effective interventions. Notably, the oral microbiota has emerged as a potential influencing factor and is increasingly becoming a focus of novel investigation in COPD research.

The human oral ecosystem contains over 700 species of microorganisms, including bacteria, viruses, and fungi, whose composition is closely linked to distinct ecological niches such as teeth, saliva, and the dorsum of the tongue [6]. Through complex interspecies interactions such as symbiosis, competition, and antagonism, these microorganisms collectively maintain oral microecological homeostasis with the host. Furthermore, the microbiota plays a significant role in modulating inflammatory signaling pathways, epigenetic modifications, immune responses, and the production of microbial metabolites, influencing the onset and progression of host diseases [7, 8]. Notably, the oral cavity and respiratory tract share a continuous anatomical pathway, providing a route for the translocation of oral microorganisms to the lungs. Oral dysbiosis or oral diseases may promote or exacerbate respiratory conditions [9, 10]. Recent observational studies have identified characteristic alterations in oral microbial diversity and core genus abundance among COPD patients across different age groups [11–13]. During acute exacerbations, reduced microbial diversity and increased microbial load are frequently observed in the lungs of affected individuals [14]. Although the oral microbiome has been implicated in COPD severity and progression, the causal relationships and underlying mechanisms require further validation.

Observational studies are often susceptible to confounding variables and reverse causality. In contrast, MR employs single-nucleotide polymorphisms (SNPs) as instrumental variables (IVs) to estimate the exposure-outcome relationship [15]. Since SNPs are randomly allocated during meiosis, with allele transmission being fixed and independent of disease status, MR analysis effectively mitigates the influence of confounding factors and reverse causation. Furthermore, bulk RNA sequencing techniques can be utilized to comprehensively elucidate the potential mechanisms of the oral microbiome in COPD pathogenesis, thus providing further validation for MR findings [16].

The current study performed a bidirectional MR analysis using summary statistics from two independent GWAS in East Asian populations, systematically evaluating the causal direction between oral microbial traits and COPD. We mapped SNPs associated with relevant microbial taxa to corresponding genes and constructed a PPI network. Eleven topological algorithms in CytoHubba were then employed to identify key hub genes. The resulting candidates were further refined using logistic regression with 1,000 repetitions of stratified 10-fold cross-validation to pinpoint core functional genes. To further validate the biological mechanisms of these genes, we integrated single-cell RNA sequencing data and performed immune infiltration analysis. Finally, molecular docking was employed to screen potential targeted therapeutics and evaluate their binding affinities. Through this multi-omics integrative approach, our study aims to elucidate the causal role and molecular basis of the oral microbiota in COPD pathogenesis, offering novel insights and potential intervention targets for disease prevention and treatment.

## 2. Materials and methods

### 2.1. Study design

This study employed a bidirectional two-sample MR approach to investigate potential causal relationships between oral microbiota and COPD. All genetic association summary statistics were derived from GWAS conducted in East Asian populations. The MR design relies on three core assumptions: 1. The genetic variants used as IVs must be strongly associated with the specific oral microbial taxa; 2. The selected SNPs should not be associated with any known or unknown confounders influencing the exposure-outcome relationship; 3. The genetic instruments influence COPD risk exclusively through their effect on oral microbiota, rather than via alternative biological pathways [15].

All analyses were performed using publicly available summary-level data. As no individual-level data were collected and no new experiments were conducted, additional ethical approval was not required. The overall study workflow is illustrated in Figure 1.

**Figure 1.**
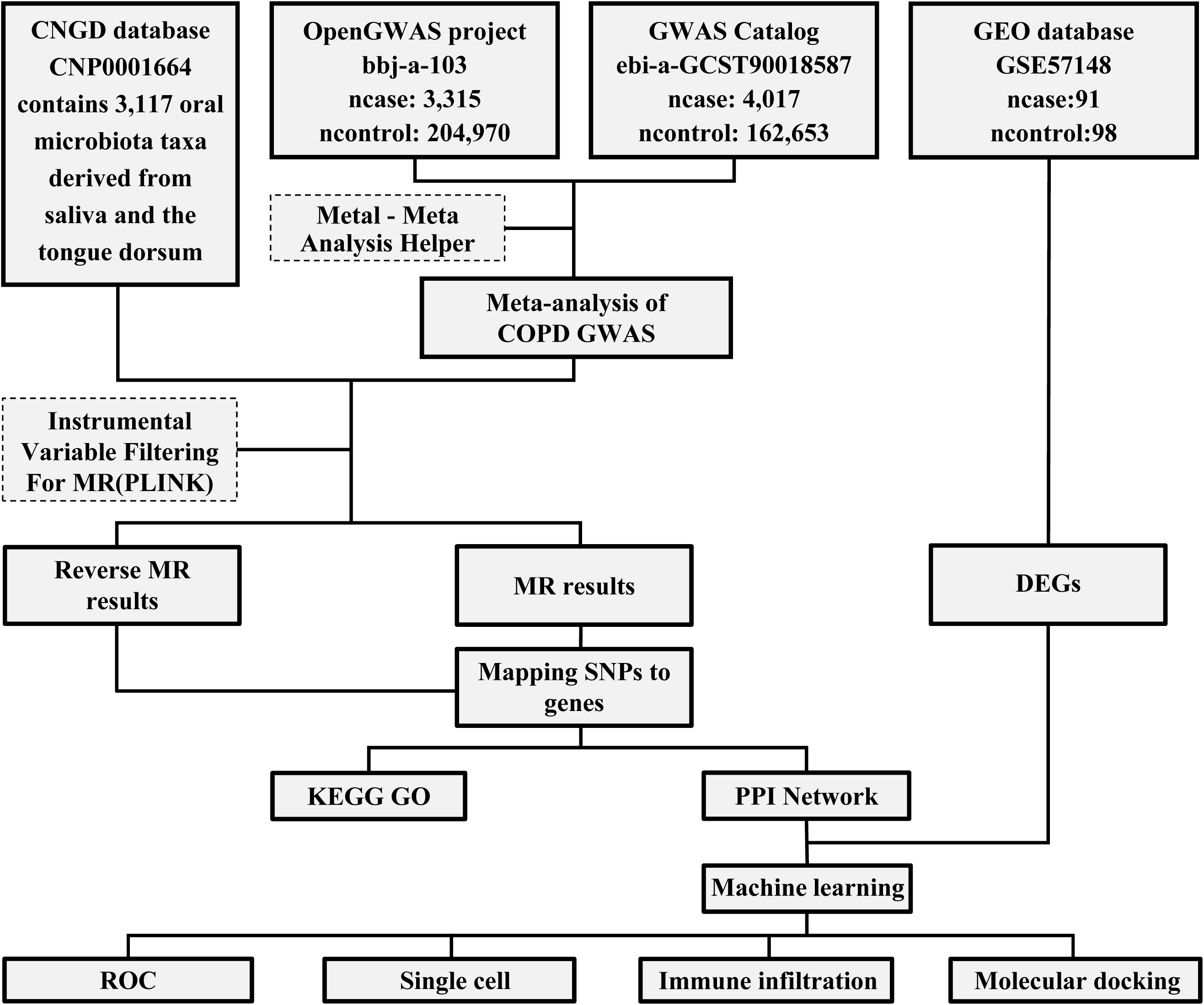
The flowchart of the study.

### 2.2. Data sources

Summary statistics for oral microbiota were obtained from a GWAS from the CNGD database, which investigated oral microbial traits in an East Asian population using 2,017 dorsum tongue samples and 1,915 saliva samples with high-depth whole-genome sequencing [17]. A comprehensive quality control pipeline was applied, including a minimum 98% variant call rate, > 20× mean sequencing depth, principal component analysis confirming no population stratification, and identity-by-descent-based exclusion of related samples. The final dataset comprised approximately 10 million genetic variants with minor allele frequency ≥ 0.5%.

Due to variations in key demographic characteristics (e.g., age, sex, living environment) across individual GWAS, which are known to affect genotype-phenotype associations, and the limited statistical power of single GWAS, we performed a meta-analysis to combine GWAS summary statistics from two independent East Asian studies. This approach enhances statistical power and reduces the likelihood of false-positive findings [18]. We incorporated COPD GWAS summary data from the OpenGWAS project and the GWAS Catalog database, comprising 7,332 cases and 364,245 controls of East Asian ancestry [19, 20]. A fixed-effects meta-analysis was conducted using the METAL software [21]. A summary of the GWAS included in this MR study is presented in Supplementary Table 1.

### 2.3. Selection of instrumental variables

In the MR analysis, we utilized the clumping procedure in PLINK to identify an adequate number of IVs while minimizing linkage disequilibrium (LD) among associated genetic variants. This approach mitigates multicollinearity arising from LD and reduces potential bias caused by weak instruments [22]. SNPs significantly associated with each oral microbial taxon were selected as IVs. Given the limited number of IVs identified under a strict genome-wide significance threshold (*p < 5×10⁻⁸*), a more inclusive criterion (*p <* 1×10⁻⁵, *r² =* 0.001, *kb =* 10,000) was applied to enhance statistical power and improve the robustness of the results. To minimize potential bias, SNPs with a minor allele frequency (MAF) below 0.01 in the original GWAS were excluded. Specific SNPs, including palindromic SNPs and those with an *F*-statistic below 10, were excluded to avoid weak IVs and reduce bias, where the F-statistic is calculated using the formula: *F = (beta/se)^2^*. Finally, Steiger filtering was conducted to retain SNPs where the exposure’s R-squared was greater than that of the outcome, ensuring the IVs did not exhibit reverse causality.

### 2.4. Bidirectional MR analysis

The study employed five complementary statistical methods to examine bidirectional causal relationships between the oral microbiome and COPD: inverse variance weighted (IVW), MR-Egger, weighted median, weighted mode, and simple mode. In analyses with multiple IVs, the IVW method served as the primary analytical approach. The IVW method combines Wald ratio estimates of individual SNPs using meta-analysis techniques to derive an overall causal estimate of the effect of oral microbiota on COPD [23]. The MR-Egger method was applied to estimate causal effects while detecting and adjusting for potential horizontal pleiotropy. The weighted median approach provides consistent causal estimates even when up to 50% of the IVs are invalid. A causal association was considered statistically significant only when the direction of effect estimates from the other four MR methods aligned consistently with that of the IVW method.

### 2.5. Sensitivity analysis

We conducted sensitivity analyses to validate the accuracy and robustness of the findings. Pleiotropy was assessed using the MR-PRESSO global test and the MR-Egger intercept test, both of which yielded *p*-values greater than 0.05, indicating no evidence of pleiotropy. Heterogeneity was evaluated with Cochran’s Q test, which also produced a *p*-value greater than 0.05, suggesting the absence of heterogeneity.

All MR analyses were conducted using R (version 4.5.3) with the TwoSampleMR and MR-PRESSO packages.

### 2.6. Mapping SNPs to genes

We utilized SNPnexus (https://www.snp-nexus.org/v4/), a web-based variant annotation tool supporting both GRCh37/hg19 and GRCh38/hg38 reference assemblies, to map each queried variant to its nearest gene, including overlapping, upstream, and downstream genes [24].

### 2.7. Functional enrichment analysis of key genes

Gene Ontology (GO) and Kyoto Encyclopedia of Genes and Genomes (KEGG) enrichment analyses were performed to explore the biological functions of COPD-related genes. GO analysis encompassed biological process (BP), cellular component (CC), and molecular function (MF) categories. These analyses were conducted using the Bioinformatics platform (https://www.bioinformatics.com.cn/).

### 2.8 Construction of PPI network

The PPI network for potential shared driver genes was generated using the STRING database (https://string-db.org/) with a medium confidence threshold (0.400). Disconnected nodes were excluded from the network, which was subsequently constructed and visualized in Cytoscape. Hub genes within this network were identified and ranked using the CytoHubba plugin by integrating 11 centrality algorithms: Betweenness, Closeness, Clustering Coefficient, Degree, DMNC, EPC, EcCentricity, MCC, MNC, Radiality, and Stress [25]. The top 30 genes from this aggregated ranking were selected. An UpSet plot was applied to pinpoint the genes consistently ranked among the top candidates by all algorithms, thereby confirming their robust centrality.

### 2.9. Bulk RNA analysis

Gene expression data were retrieved from the GSE57148 dataset in the GEO database, comprising 91 controls and 98 COPD patients of East Asian descent [26]. After initial data processing, differential expression analysis was performed using the limma package. Genes were deemed significant if |logFC| > 0.25 and *p* < 0.05, ensuring biological relevance and false positive control [27]. Subsequently, expression heatmaps were generated based on these genes.

Genes identified through CytoHubba screening were then intersected with differentially expressed genes (DEGs) from the bulk transcriptome, and the overlapping candidates were further refined through logistic regression.

### 2.10. Logistic regression

To identify key genes associated with COPD driven by oral microbiota, we fitted univariable and multivariable logistic regression models on the candidate genes and used DeLong’s test for correlated ROC curves to retain the most parsimonious model. The robustness of the selected model was then evaluated through 1,000 repetitions of stratified 10-fold cross-validation and bootstrap-based internal validation with optimism correction, with the area under the receiver operating characteristic curve (AUC) quantifying its discriminative performance.

### 2.11. Single-cell analysis

Lung tissue samples from five East Asian COPD patients and three non-smokers were obtained from the GEO database under accession number GSE173896 [28]. The Seurat package initially filtered scRNA-seq data to obtain higher-quality cells, retaining cells with between 200 and 4,500 detected genes, total UMI counts between 1,000 and 35,000, and a mitochondrial transcript percentage below 10%. After quality control and data filtering, PCA was performed on the top 2,000 highly variable genes, followed by Harmony-based batch correction across groups. Utilizing UMAP, we achieved unsupervised clustering and unbiased visualization of cell subpopulations. The lung tissue module within the scMayoMap package was utilized to annotate subpopulations within each cluster [29].

Additionally, single-cell differential expression analysis was applied to annotated lung tissue cells, enabling a direct comparison of target gene expression between COPD patients and healthy controls.

### 2.12 Immune infiltration

The CIBERSORT algorithm was utilized to estimate the relative proportions of 22 immune cell types in each sample group. Pearson correlation analysis was conducted to examine the associations between key genes and immune cell subtypes.

### 2.13. Molecular docking analysis

Drug-gene interactions were explored using DSigDB (https://dsigdb.tanlab.org/DSigDBv1.0/). Drug structures were obtained from the PubChem database (https://pubchem.ncbi.nlm.nih.gov), while protein structures of the candidate genes were retrieved from the Protein Data Bank (http://www.rcsb.org/). CB-Dock2 (https://cadd.labshare.cn/cb-dock2/index.php) was used for protein-ligand blind docking, and the cavity size and Vina score were evaluated.

## 3. Results

### 3.1. Causal impact of the oral microbiome on COPD development

Figure 2 presents the results of the correlation analysis between genetically predicted oral microbiota and COPD risk. Among 3,117 bacterial taxa examined in the forward MR analysis, 48 oral microbial features demonstrated statistically significant associations with COPD. Of these, 24 taxa (15 from the dorsum tongue and 9 from saliva) exhibited protective effects (OR < 1), while 24 taxa (11 from the dorsum tongue and 13 from saliva) were identified as risk factors (OR > 1). These microorganisms were primarily concentrated within the genera *Fusobacterium*, *Neisseria*, *Pauljensenia*, *Prevotella*, and *Streptococcus* (Figure 2A). In the reverse MR analysis, 79 microbial taxa were identified as having potential reverse causal relationships with COPD. Among these, 37 features (25 from the dorsum tongue and 12 from saliva) showed protective effects (OR < 1), whereas 42 taxa (16 from the dorsum tongue and 26 from saliva) were classified as risk factors (OR > 1). These taxa were predominantly enriched in the genera *Campylobacter_A*, *Rothia*, and *Streptococcus* (Figure 2B). A Sankey diagram illustrating the distribution of microbial taxa at the order, family, and genus levels is presented in Figure 2C.

**Figure 2.**
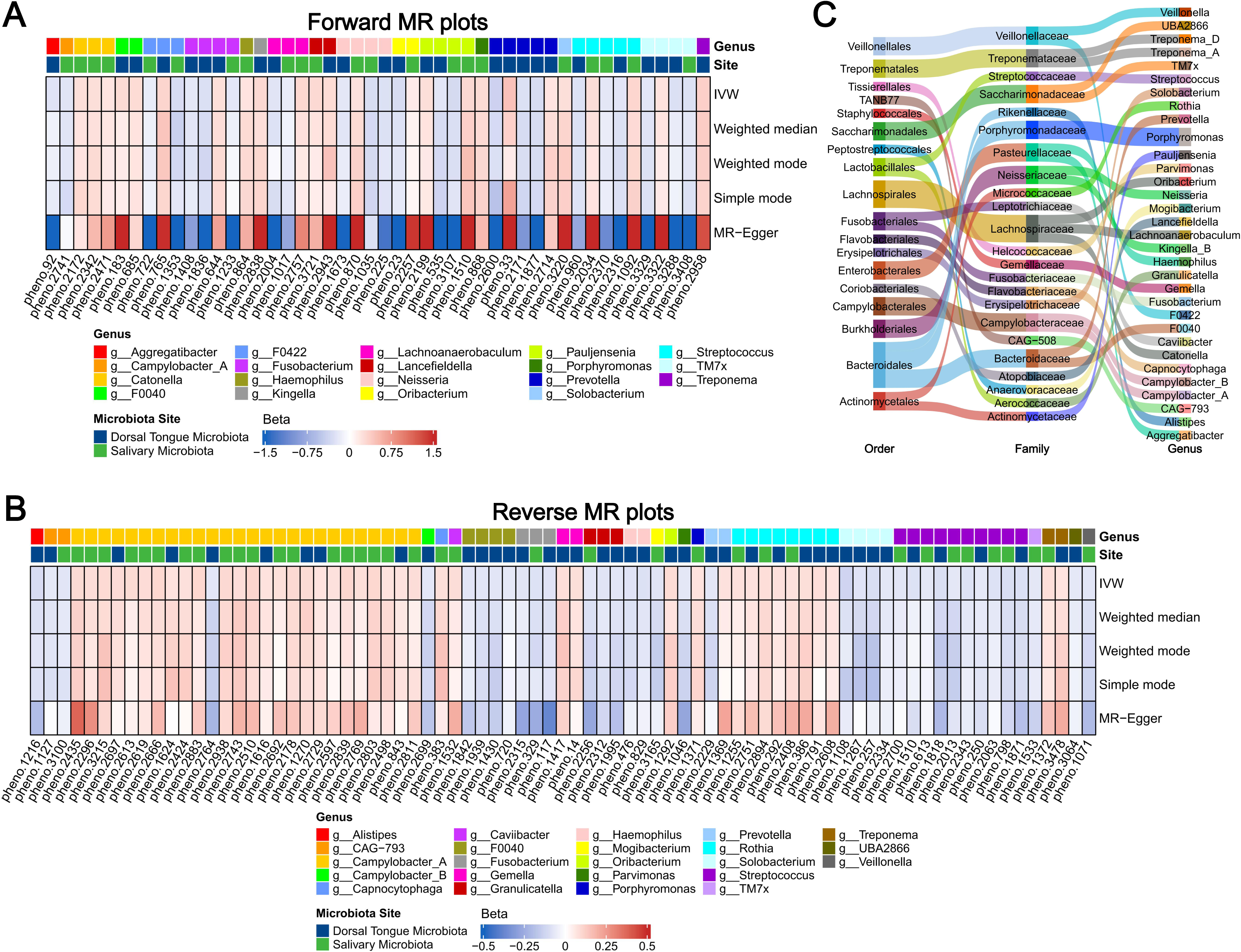
Species-level oral microbiota causally associated with COPD identified by bidirectional MR analysis. (A) Dorsal tongue and salivary species with forward causal effects on COPD risk. (B) Dorsal tongue and salivary species with reverse causal effects influenced by COPD. (C) Sankey plot illustrating the taxonomic relationships among causally associated taxa at the order, family, and genus levels.

Furthermore, all five MR methods yielded consistent directional effects and statistical significance. The *F*-statistics for all selected SNPs exceeded 10, ruling out weak instrument bias. The MR-Egger regression intercept did not deviate significantly from zero (*p* > 0.05), and the MR-PRESSO global test detected no outliers, indicating no evidence of horizontal pleiotropy. Cochran’s Q test revealed no substantial heterogeneity (*p* > 0.05). The Steiger directionality test ruled out reverse causation (Supplementary Table 2; Supplementary Table 3).

### 3.2. Genes and functionality

We performed functional annotation on genes mapped by SNPs associated with oral microbiota-related COPD to elucidate their potential biological functions (Supplementary Table 4). KEGG pathway enrichment analysis (Figure 3A) revealed that the key genes were associated with cell adhesion molecules, bacterial invasion of epithelial cells, Th1 and Th2 cell differentiation, and focal adhesion. For BP (Figure 3B), these genes were primarily involved in cell-cell adhesion via plasma-membrane adhesion molecules and cell junction assembly. CC analysis (Figure 3C) indicated enrichment in ion channel complex, transporter complex, and lamellipodium. MF analysis (Figure 3D) showed enrichment in transmembrane transporter binding, gated channel activity, and cell-cell adhesion mediator activity (Supplementary Table 5).

**Figure 3.**
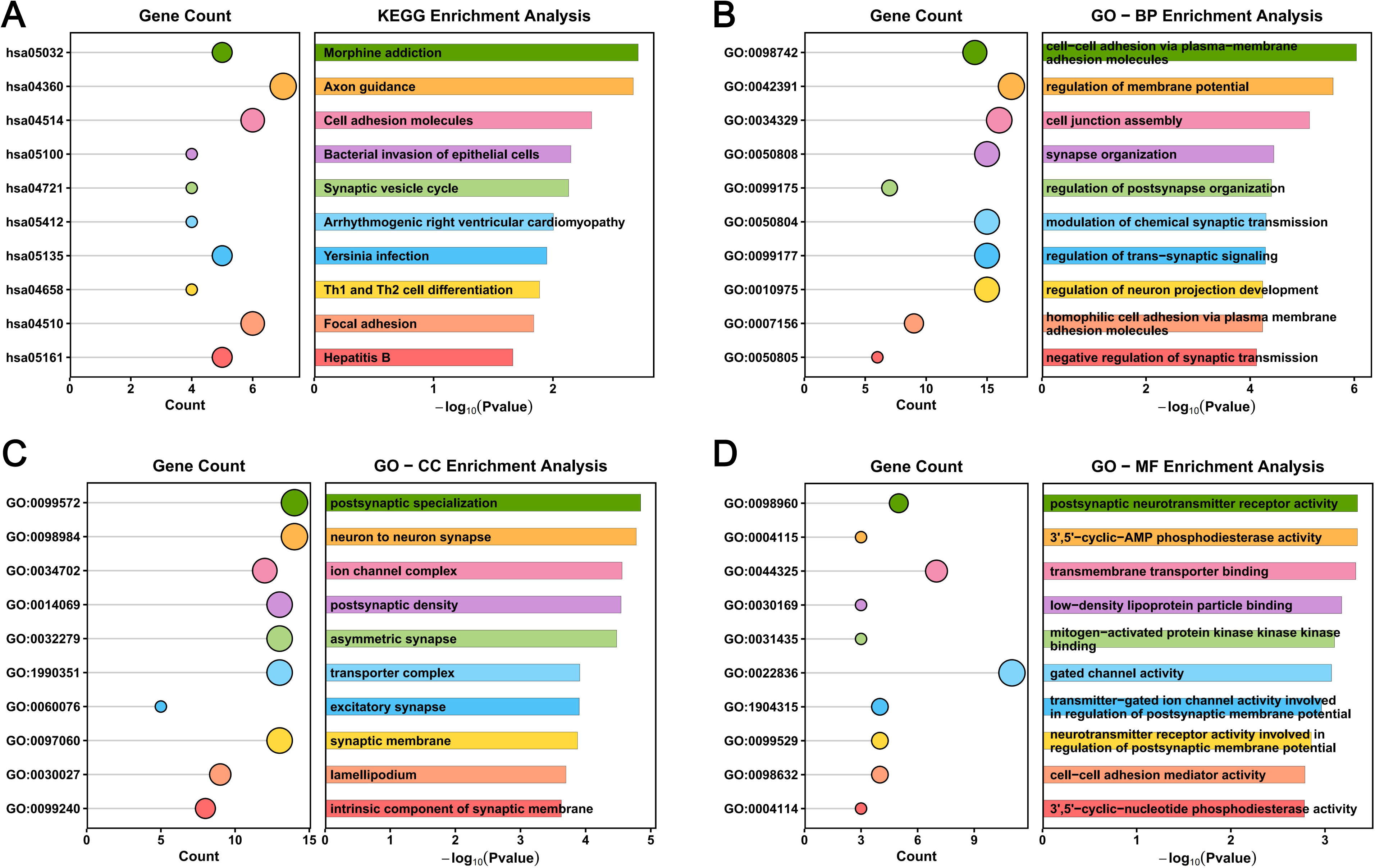
Functional enrichment analysis of DEGs associated with COPD. (A) KEGG pathway enrichment analysis. (B) GO biological process (BP) enrichment analysis. (C) GO cellular component (CC) enrichment analysis. (D) GO molecular function (MF) enrichment analysis.

### 3.3. Identification of PPI network

From the 344 shared driver genes identified through SNP mapping, following the exclusion of disconnected nodes, hub genes were ranked by 11 centrality algorithms implemented in CytoHubba. An UpSet plot subsequently highlighted 10 genes that ranked consistently among the top 30 across all 11 algorithms (Figure 4).

**Figure 4.**
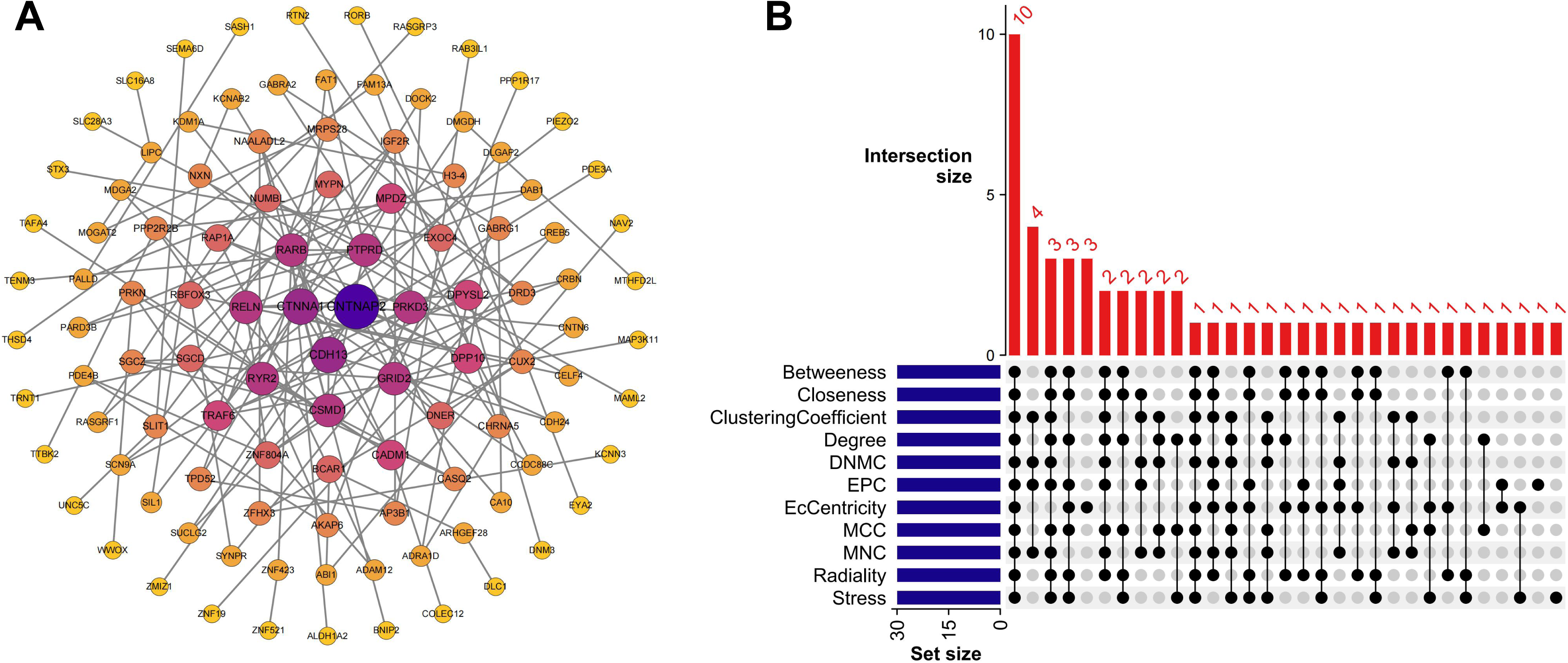
PPI network analysis and hub gene identification. (A) PPI network constructed via Cytoscape with isolated nodes removed. (B) Upset plot showing the intersection of the top 30 genes ranked by 11 CytoHubba algorithms.

### 3.4. Identification of DEGs in an East Asian dataset

In the GSE57148 dataset, comprising COPD patients and healthy controls of East Asian descent, 2,814 (DEGs) were identified, including 1,326 up-regulated and 1,488 down-regulated genes (Figure 5). Among these, *CDH13* and *MPDZ* were identified as overlapping genes between the CytoHubba hub genes and the bulk RNA-seq DEGs.

**Figure 5.**
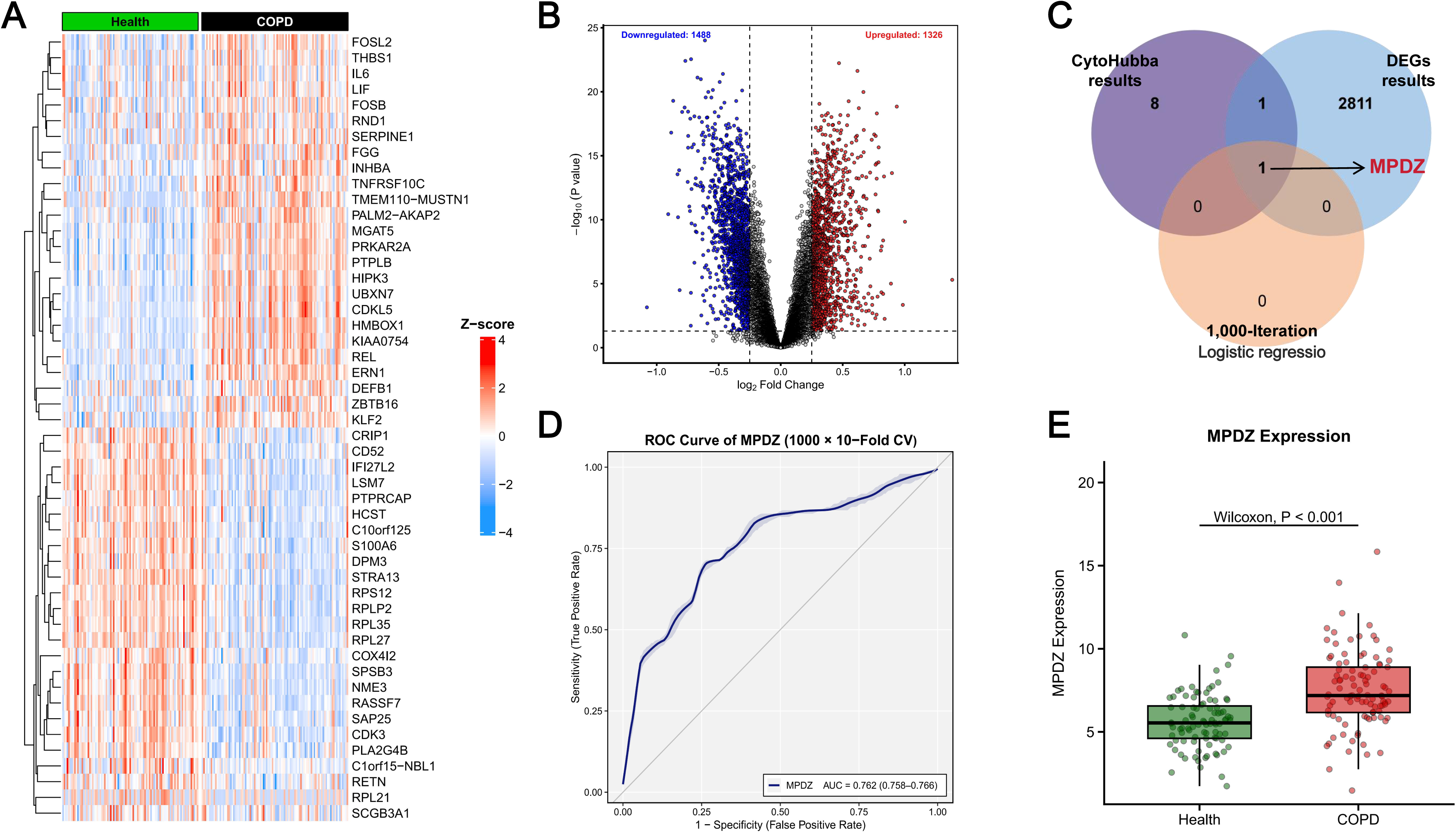
Identification and validation of the hub gene *MPDZ*. (A) Clustering heatmap of GSE57148. (B) Volcano plot of GSE57148. (C) Venn diagram showing the intersection of CytoHubba-derived genes, DEGs from GSE57148, and logistic regression-selected genes from 1,000 iterations. (D) ROC curve of *MPDZ* from 1,000 rounds of 10-fold cross-validation. (E) Box plot of *MPDZ* expression between COPD patients and healthy controls.

### 3.5. Validation of core genes for COPD

In the GSE57148 dataset, MPDZ was significantly upregulated in COPD compared with healthy controls (*p* < 0.001). The DeLong test indicated that adding *CDH13* to *MPDZ* did not significantly improve discrimination (*p* = 0.562), whereas *CDH13* alone (AUC = 0.601) was significantly inferior to both models, supporting selection of the *MPDZ* single-gene model. Higher *MPDZ* expression was associated with increased COPD risk (OR = 1.705, 95% CI = 1.406 - 2.067, *p* < 0.001), with a likelihood-ratio χ² of 41.894 (p < 0.001) and a Nagelkerke pseudo R² of 0.265.

Across 1,000 repetitions of stratified 10-fold cross-validation, the mean AUC was 0.762 (95% CI = 0.758 - 0.766). Optimism-corrected internal validation based on 200 bootstrap resamples yielded a corrected C-statistic of 0.770, a calibration slope of 1.033, and a calibration intercept of 0.015, with a mean absolute calibration error of 0.036 between predicted and observed probabilities (Figure 5).

### 3.6. Results of single-cell analysis

We analyzed scRNA-seq data from 8 samples, comprising lung cells from three healthy controls and five COPD patients. After stringent quality control, 13 distinct cell clusters were identified and annotated using the lung tissue module of the scMayoMap package. Cell types included B cells, Basal cells, Capillary aerocytes, Ciliated cells, Endothelial cells, Epithelial cells, Fibroblasts, Lymphoid cells, Macrophages, Mesothelial cells, Neutrophils, Pulmonary alveolar type II cells, and T cells (Figure 6). Single-cell differential expression analysis revealed that *MPDZ* was significantly upregulated in Ciliated cells from COPD patients compared to healthy individuals (Supplementary Table 6).

**Figure 6.**
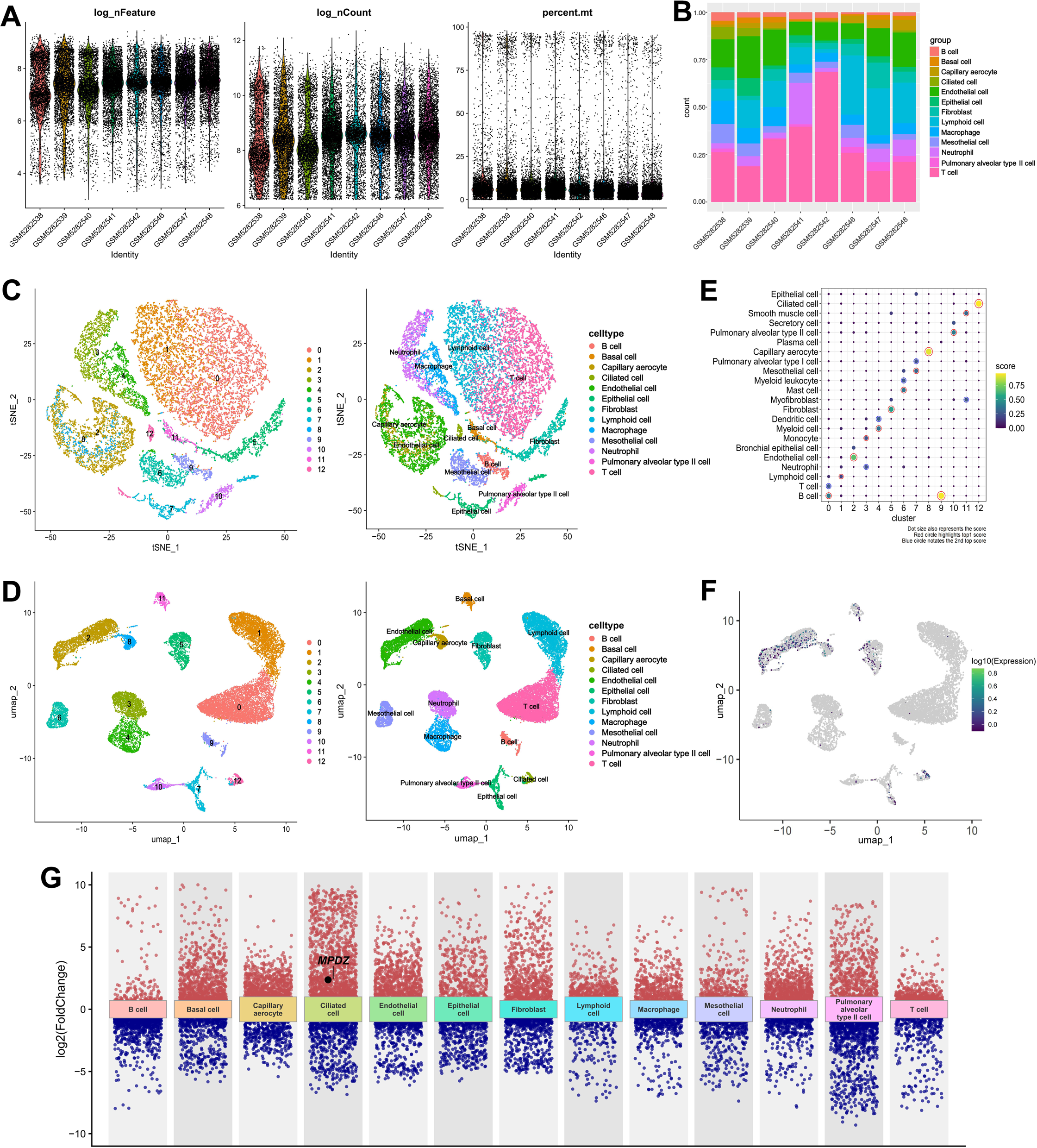
Single-cell RNA sequencing analysis and cell-type-specific localization of MPDZ. (A) Violin plots of quality control metrics including log-transformed feature count, UMI count, and mitochondrial gene percentage. (B) Stacked bar plot showing cell type composition across different samples. (C) t-SNE visualization of 13 cell clusters annotated by scMayoMap. (D) UMAP visualization of 13 cell clusters annotated by scMayoMap. (E) Annotation confidence scores assigned by scMayoMap for lung tissue cell types. (F) Expression distribution of *MPDZ* across cell clusters. (G) Cell-type-specific differential expression landscape, with each dot representing a gene plotted by log2 (FoldChange); *MPDZ* is highlighted in ciliated cells.

### 3.7. Immuno-infiltration analysis by CIBERSORT

Cell type proportions were estimated using the CIBERSORT algorithm. Compared with normal samples, COPD samples showed significant differences in the proportions of four immune cell types: Eosinophils, Neutrophils, resting NK cells, and T follicular helper cells. Correlation analysis revealed that *MPDZ* was significantly positively correlated with activated Dendritic cells, resting Mast cells, Monocytes, Neutrophils, and resting memory CD4 T cells, but negatively correlated with memory B cells, resting Dendritic cells, regulatory T cells (Tregs), follicular helper T cells, M1 Macrophages, and M2 Macrophages in COPD (Figure 7).

**Figure 7.**
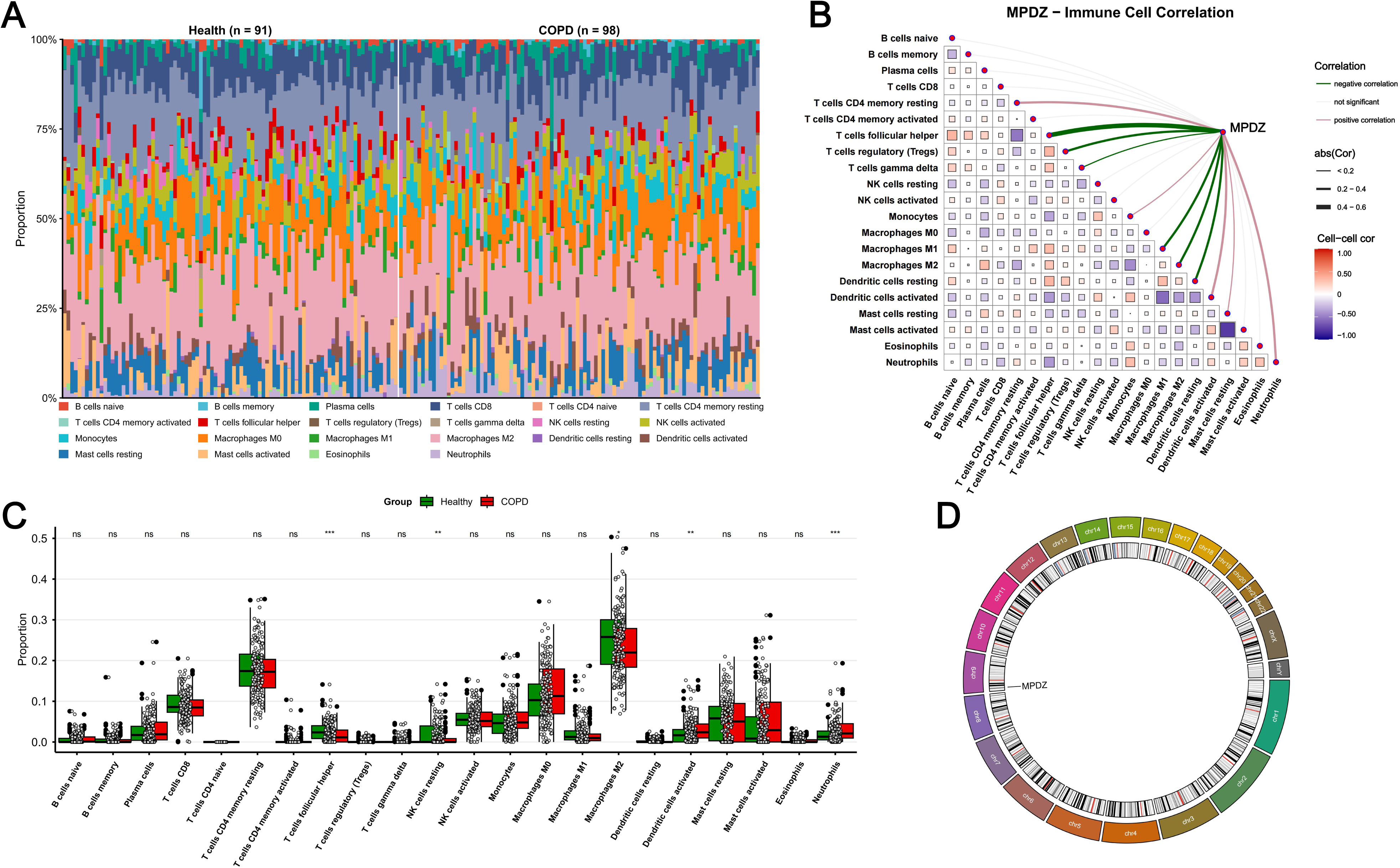
Immune infiltration analysis and correlation with *MPDZ* expression. (A) CIBERSORT-estimated immune cell proportions in healthy controls and COPD patients. (B) Correlation between *MPDZ* expression and 22 immune cell types. (C) Comparison of immune cell proportions between healthy controls and COPD patients. ns, not significant; \**P* < 0.05; \*\**P* < 0.01; \*\*\**P* < 0.005. (D) Chromosomal localization of *MPDZ*.

### 3.8. Molecular docking evaluation

Based on the above results, six active compounds were selected for molecular docking with *MPDZ*: Captopril, -5.7 kcal/mol; Camptothecin, -8.0 kcal/mol; Digitoxin, -9.9 kcal/mol; Irinotecan, -8.7 kcal/mol; Pregnenolone, -7.3 kcal/mol; and Trichostatin A, -7.3 kcal/mol. The optimal docking conformations are visualized in Figure 8. All binding energies were < -5 kcal/mol, indicating strong binding affinities.

**Figure 8.**
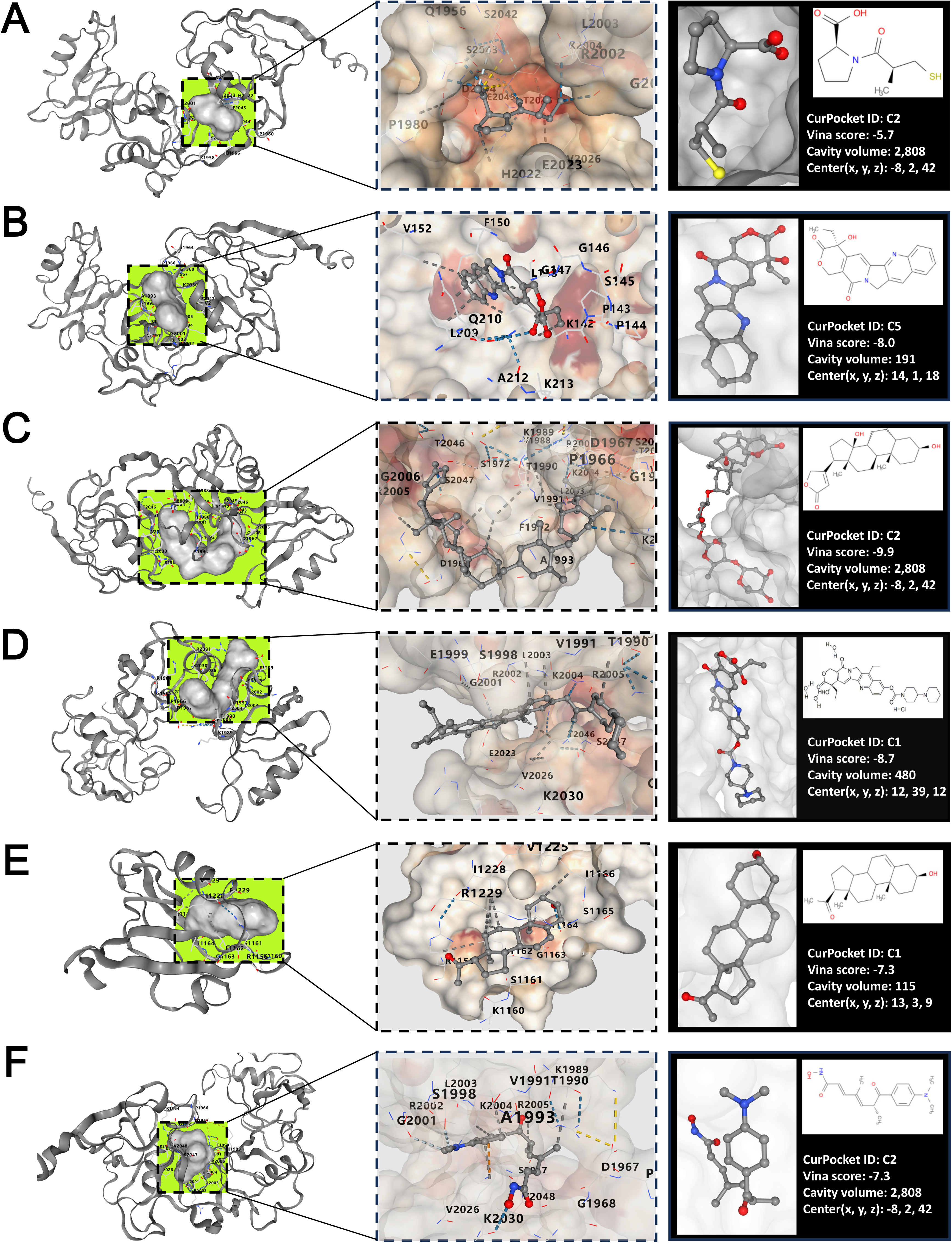
In silico molecular docking of MPDZ with (A) captopril, (B) camptothecin, (C) digitoxigenin, (D) irinotecan, (E) pregnenolone, and (F) trichostatin A.

## 4. Conclusion

Compared with the extensively studied gut microbiome, research on the oral microbiome in COPD remains relatively limited. Observational studies are often affected by technical and sampling variability, producing heterogeneous findings that obscure the underlying pathogenesis. We employed bidirectional MR to investigate causal associations between the oral microbiome and COPD, focusing on the biological mechanisms by which oral microbiota may contribute to COPD pathogenesis and on the identification of candidate effector genes. By integrating bulk and single-cell transcriptomic data with logistic regression, we prioritized candidate genes potentially relevant to COPD pathogenesis. Drug prediction and molecular docking analyses further suggested in silico binding partners for the top candidate. Through this multi-omics framework, our study provides genetic evidence linking the oral microbiota to COPD and identifies candidate effector genes for future mechanistic investigation.

This study performed a bidirectional MR analysis using oral microbiome data from the CNGD database and a meta-analyzed COPD GWAS dataset that integrated East Asian population statistics from both the GWAS Catalog and OpenGWAS project through METAL. The MR results identified significant microbial taxa primarily within the genera *Fusobacterium*, *Neisseria*, *Pauljensenia*, *Prevotella*, and *Streptococcus*, with notable heterogeneity observed at the species level. The pathogenesis of COPD has been closely associated with dysregulation of pro-inflammatory cytokines. *Streptococcus* polysaccharide capsule reduces the effectiveness of host defense mechanisms, which normally cause its clearance [30]. However, the death of *pneumococci*, whether through host-mediated killing or autolysis, not only results in bacterial clearance but also in the release of inflammatory agents such as the pore-forming toxin, pneumolysin, and bacterial cell wall components that intensify local inflammation [31, 32]. While substantial evidence has established *Streptococcus* as a risk factor for COPD, emerging studies indicate that specific streptococcal species may exert protective effects. For instance, pulmonary *Streptococcus sanguinis* has been shown to suppress NF-κB pathway activation and inhibit IL-8 production triggered by lipopolysaccharide or H₂O₂, attenuating inflammation and oxidative stress in COPD [33]. Similarly, oral *Streptococcus oxalis* activates innate immunity and reduces influenza-induced pulmonary inflammation [34]. The heterogeneous associations observed for *Streptococcus* in our study align with this evolving understanding, reinforcing the notion that microbial functions are often species-specific. In contrast, *Prevotella* is generally less frequently implicated as a risk factor for respiratory diseases, with only limited strains demonstrating opportunistic pathogenic potential [35]. Accumulating evidence reveals that *Prevotella* abundance was inversely related to COPD severity in terms of symptoms and positively related to lung function and exercise capacity [36]. Mechanistically, *Prevotella* contributes to the process of airway epithelial gene expression and suppresses inflammatory signaling pathways. Furthermore, within the respiratory microbial ecosystem, *Prevotella* exhibits a low pro-inflammatory immunophenotype. In healthy individuals, it collaborates with other commensal bacteria to induce immune tolerance, facilitating microbial colonization and maintaining mucosal homeostasis [37]. Regarding *Fusobacterium*, existing research presents seemingly contradictory findings. On one hand, *Fusobacterium. nucleatum* (*F. n*) has been identified as a pathogen that induces the production of IL-6 and IL-8, exacerbating respiratory diseases, especially COPD [38, 39]. On the other hand, *F. n* infection has been shown to alleviate aortic atherosclerosis symptoms in animal models [40]. *Pauljensenia* has been associated with reduced risks of bronchitis and tonsillitis, though the underlying mechanisms remain unclear [41]. However, as a newly established genus, *Pauljensenia* has received limited attention in the context of COPD, and its mechanistic role in COPD pathogenesis requires further validation. At the species level, the relationships between most bacterial taxa and disease states remain insufficiently investigated. The complexity of oral microbial communities and substantial inter-individual variability present significant challenges for research. Therefore, further studies are essential to elucidate the role of oral bacteria in respiratory diseases such as COPD, which will facilitate the development of potential therapeutic strategies.

Furthermore, the reverse MR analysis identified potential causal effects of COPD on specific oral microbial taxa. Our findings identified significant associations primarily involving *Rothia*, *Campylobacter_A*, and *Streptococcus*. Specifically, COPD appears to promote the proliferation of *Campylobacter_A* and *Rothia*, while simultaneously suppressing *Streptococcus* abundance. These results align with existing research evidence. Multiple cohort studies have demonstrated a significant increase in the relative abundance of *Campylobacter* among COPD patients, with its levels positively correlating with metabolic characteristics of eosinophilic COPD [42, 43]. Time-series analyses further confirm that fluctuations in *Campylobacter* abundance closely parallel dynamic changes in inflammatory markers [44]. In COPD patients with HIV co-infection, *Campylobacter* abundance similarly shows marked elevation, suggesting that compromised immune function and inflammation collectively promote the ecological advantage of this bacterium [45]. *Rothia*, as a common oral-derived bacterium, has been observed to be significantly enriched in lung samples from COPD patients and associated with disease stability or inflammatory indicators [45]. Specifically, its relative abundance increases 2.38-fold in COPD patients compared to healthy controls and shows positive correlations with cytokines such as IL-4 and IL-5, indicating that the inflammatory microenvironment in COPD drives ecological niche expansion of *Rothia* and its potential beneficial roles [46, 47]. Multiple studies have reported reduced abundance of *Streptococcus* in both oral and respiratory tracts of COPD patients, potentially attributable to enhanced inflammation, increased antimicrobial peptide production, or therapeutic interventions. This suppression appears more pronounced during stable disease phases and in high-risk patients [48, 49].

To characterize the biological functions of candidate genes, we mapped MR-identified IVs to their genomic loci and performed GO and KEGG enrichment analyses. KEGG analysis identified enrichment in cell adhesion molecules and focal adhesion, both central to airway epithelial-matrix interactions. FAK activation by the COPD-associated glycoprotein YKL-40 drives smooth muscle proliferation and fibrotic remodeling via the MAPK cascade [50, 51]. The enrichment of Th1 and Th2 cell differentiation captures the adaptive immune dysregulation in COPD, where NLRP3-mediated pyroptosis triggers Th1-skewed IFN-γ production, while Th2-derived IL-4 and IL-13 promote mucus hypersecretion and airway hyperresponsiveness [52, 53]. Bacterial invasion of epithelial cells is directly relevant to the oral microbiota context of this study, given that oral-derived anaerobes reach the lower airways through microaspiration, where their cysteine proteases disrupt the protease-antiprotease balance and sustain chronic inflammation [54, 55]. Refining these findings, BP and CC terms localized these impairments to plasma-membrane adhesion complexes and ion channel machinery. In the COPD airway, cigarette smoke and microbial products compromise adherens and tight junction integrity by epigenetically silencing the *CDH1* enhancer and disrupting zona occludens-1 through aberrant phosphorylation, increasing paracellular permeability and impairing epithelial regeneration [56, 57]. CC enrichment in ion channel complex and transporter complex corresponds to acquired CFTR and BK channel dysfunction, where TGF-β1-mediated transcriptional suppression depletes airway surface liquid and creates a permissive niche for bacterial colonization [58, 59], while enrichment of lamellipodium reflects Rho GTPase-dependent cytoskeletal reorganization that facilitates inflammatory cell infiltration and fibroblast migration during airway remodeling [60]. MF analysis corroborated these observations, with transmembrane transporter binding and gated channel activity reinforcing the ion transport defect, and cell-cell adhesion mediator activity echoing the junctional impairment identified at the BP level. These multi-level enrichments indicate that oral microbiota-associated genetic variants may predispose to COPD through a cascade from epithelial barrier compromise and mucociliary dysfunction to dysregulated immune responses against microbial challenge.

Among the CytoHubba hub genes intersected with DEGs from the East Asian GSE57148 dataset, *CDH13* and *MPDZ* were identified as overlapping candidates, with *MPDZ* consistently selected by logistic regression across 1,000 iterations. *CDH13* encodes T-cadherin, a GPI-anchored adiponectin receptor whose variants are linked to plasma adiponectin levels and COPD risk in Chinese populations [61]. Genome-wide interaction studies further indicate that *CDH13* polymorphisms modify the effect of particulate matter on lung function decline in East Asian and European cohorts [62, 63]. *MPDZ* is a multi-PDZ domain scaffolding protein that stabilizes tight junctions by linking claudins and JAM proteins to intracellular signaling. Its upregulation in COPD ciliated cells may represent a compensatory response to chronic junctional disruption. In lung cancer, MPDZ is silenced through promoter hypermethylation, and its loss promotes metastasis via Hippo-YAP dysregulation [64]. The sustained epithelial injury in advanced COPD may progressively exhaust this compensatory capacity, converging with the *MPDZ* loss observed in malignancy and suggesting a shared vulnerability in tight junction integrity that underlies the epidemiological link between these two conditions.

Drug prediction analysis identified captopril, camptothecin, digitoxigenin, irinotecan, pregnenolone, and trichostatin A as candidate therapeutics targeting *MPDZ*. Captopril, an ACE inhibitor, has shown anti-inflammatory effects in COPD models [65]. Camptothecin analogues exhibit anti-inflammatory properties in pulmonary diseases [66]. Digitoxigenin, through its modulation of ion channels, may alleviate COPD symptoms [67]. Irinotecan, a topoisomerase inhibitor, potentially suppresses fibrotic pathways [68]. Pregnenolone supports steroidogenesis and counter hormonal imbalances in COPD [69]. Trichostatin A, an HDAC inhibitor, restores diminished HDAC activity in COPD and enhances anti-inflammatory responses [70]. Molecular docking confirmed strong binding affinities for these compounds, suggesting their therapeutic potential.

Nevertheless, several limitations should be acknowledged. The composition of the oral microbiome is influenced by multiple factors, including genetic background, dietary habits, lifestyle, and environmental exposures, all of which could affect the observed associations. Additionally, the MR analysis focused on East Asian populations, and the findings may not be directly generalizable to other ethnic groups. Further studies are warranted to validate and extend these results across diverse populations. The quality of scRNA-seq data is also highly dependent on technical factors such as cell capture efficiency and sequencing depth, which may lead to under-representation of certain cell types. In the field of drug prediction, although DSigDB provides extensive gene-drug association data, these predictions require experimental validation to confirm efficacy and safety. Furthermore, drug effects observed at the cellular level may not directly translate to in vivo outcomes, and clinical trials are necessary to verify practical applicability.

In summary, this study used bidirectional MR analysis to identify causal associations between oral microbial taxa and COPD in East Asian populations. SNPnexus was used to map COPD-associated genes from the relevant SNPs, linking microbial exposure to candidate effector genes. Single-cell analysis of COPD lung tissue revealed cell-type-specific expression patterns of these genes, and bulk RNA sequencing identified distinct expression profiles between COPD patients and controls. Through PPI network analysis, logistic regression, and molecular docking, MPDZ was prioritized as a candidate gene with several in silico-predicted small-molecule interactions. Collectively, these findings prioritize candidate targets relevant to COPD pathogenesis and provide a hypothesis-generating basis for future mechanistic studies and therapeutic exploration.

## 5. Conclusion

This study presents a comprehensive bioinformatics approach to evaluate the causal relationship between oral microbiota and COPD. Our analysis identified 48 oral microbial taxa with forward causal effects on COPD and 79 taxa demonstrating reverse causal relationships. Through the integration of 11 topological algorithms in CytoHubba, transcriptomic differential expression analysis, logistic regression validation, single-cell profiling, immune infiltration assessment, and molecular docking, *MPDZ* was identified as a potential key regulator in COPD-related immune modulation. These findings offer candidate targets and a theoretical basis for future investigations into COPD pathogenesis and therapeutic development.

## Author contributions

Xiao-Jie An: Conceptualization, Data curation, Formal analysis, Methodology, Project administration, Software, Supervision, Validation, Visualization, Writing - original draft, Writing - review and editing.

Zi-Feng Wei: Conceptualization, Data curation, Formal analysis, Methodology, Project administration, Software, Supervision, Validation, Visualization, Writing - original draft, Writing - review and editing.

Yi-Ting Huang: Data curation, Methodology, Supervision, and Visualization. Jia-Pei Wuzhang: Writing - original draft, Writing - review and editing.

Xin-Xin Zhang: Data curation Hao-Yu Li: Data curation Run-Ze Liu: Validation

## Data availability

The GWAS summary data for COPD analyzed in the current study are sourced from bbj-a-103 and GCST90018587 (OpenGWAS project: https://opengwas.io/datasets/; GWAS Catalog: https://www.ebi.ac.uk/gwas/). The GWAS summary data for salivary and dorsal tongue microbiota are sourced from CNP0001664 (CNGB database: https://ftp2.cngb.org/). The RNA bulk and single-cell data for COPD are sourced from GSE57148 and GSE173896 (GEO database: https://www.ncbi.nlm.nih.gov/geo/).

## Declarations

### Ethics statement

The present study does not require ethical approval.

